# Eligibility for hepatitis B virus (HBV) treatment and prevalence of drug resistance in a Ugandan population cohort

**DOI:** 10.1101/2025.04.06.25325341

**Authors:** Sheila F Lumley, Beatrice Kimono, Joseph Mugisha, Ronald Makanga, Moses Kwizera Mbonye, Kevin Ojambo, Elizabeth Waddilove, Chris Kent, Brian Ssengendo, Richard Ndungutse, Anna L McNaughton, Florence Nambaziira Muzaale, Janet Seeley, Josh Quick, Ponsiano Ocama, Robert Newton, Philippa C Matthews

**Affiliations:** Peter Medawar Building for Pathogen Research, Nuffield Department of Medicine, University of Oxford, South Parks Road, Oxford, UK; Department of Infectious Diseases and Microbiology, Oxford University Hospitals NHS Foundation Trust, UK; MRC/UVRI and LSHTM Uganda Research Unit, Uganda; The Francis Crick Institute, 1 Midland Road, London, UK; Institute of Microbiology and Infection, University of Birmingham, UK; Population Health Sciences, Bristol Medical School, University of Bristol, UK; London School of Hygiene and Tropical Medicine, London, UK; Makerere University College of Health Sciences in Kampala, Uganda; Department of Health Sciences, University of York, UK; Division of Infection and Immunity, University College London, Gower St, London, UK; Department of Infection, University College London Hospitals, 235 Euston Rd, London, UK

**Keywords:** HBV, Uganda, treatment, entecavir, tenofovir, liver function, APRI, elastography

## Abstract

**Background:** The World Health Organization (WHO) 2024 Hepatitis B virus (HBV) treatment guidelines expand eligibility for nucleos(t)ide analogue treatment in individuals with chronic HBV infection. For countries to implement these guidelines, there is a critical need to understand the population who are treatment eligible. While HBV drug resistance (HBVDR) is uncommon, monitoring for any potential resistance is a relevant public health consideration.

**Methods:** We studied a population in rural Uganda to describe the proportion of individuals eligible for treatment based on the 2024 WHO treatment guidelines. We determined how this proportion varies according to the eligibility criteria used, comparing the performance of different assessment tools. We calculated Aspartate Aminotransferase-to-Platelet Ratio Index; APRI, Gamma-Glutamyl Transpeptidase-to-Platelet Ratio; GPR and transient elastography; TE and performed HBV sequencing using Oxford Nanopore Technology to determine the prevalence of HBVDR in treatment naive and treatment experienced individuals.

**Results:** In this cohort, 24/63 (38%) individuals with CHB were eligible for treatment. This fell to 14/63 (22%) in a hypothetical scenario where TE was not available for the assessment of liver fibrosis. We demonstrate a lack of concordance between non-invasive tests (NIT) of liver disease in treatment-naive HBV mono-infected individuals. An APRI cut-off of 0.5 had a sensitivity of 23.0% for predicting a TE score of >7 kPa (F2 fibrosis). Sensitivity for detecting F2 fibrosis was increased to 38.5% using an APRI cut off of 0.36, and to 46.2% using the GPR. We did not identify any HBVDR in the HBV mono-infected treatment-naive population (n=58). 24/210 individuals were living with HIV/HBV coinfection; HBV was sequenced in 5 of these of whom 2 had genomic evidence of nucleos(t)ide analogue resistance (rt180M/204V).

**Conclusions:** While the WHO 2024 treatment criteria offer an opportunity to expand access to care, there is a need to determine how assessment tools differ in determination of eligibility in different settings. HBVDR remains uncommon but more research is needed to understand its prevalence and clinical impact in African populations.

## 1. Introduction

The World Health Organisation (WHO) African region has among the highest incidence and mortality rates from liver disease in the world due to the high prevalence of viral hepatitis, alcohol intake, and metabolic dysfunction associated steatotic liver disease (MASLD)^1^. Hepatitis B virus (HBV) infection is a significant public health challenge in this region. Despite the availability of safe and effective HBV vaccines and nucleos(t)ide analogue therapy, an estimated 65 million people are living with chronic HBV (CHB) infection, and approximately 270,000 HBV-related deaths occur in Africa each year. Access to effective antiviral treatments which can reduce transmission, cirrhosis, HCC and improve long term survival remains limited in this region; at the end of 2022, only 4% of people living with CHB had been diagnosed and 0.2% had received antiviral therapy^2^. In 2024, the WHO published new expanded and simplified HBV treatment guidelines aiming to accelerate progress towards HBV elimination^3–5^. For countries to implement these guidelines, there is a critical need to understand the population which is treatment eligible. While HBV drug resistance (HBVDR) is uncommon, monitoring for any potential resistance is a relevant public health consideration.

The 2024 WHO HBV treatment guidelines^6^ present four options for determining treatment eligibility based on the presence of liver fibrosis/cirrhosis (APRI or TE), hepatic inflammation (alanine aminotransferase - ALT), HBV replication (HBV VL) and past medical and family history. The guidance provides simplified criteria for use when access to laboratory infrastructure and specialised tests is limited (for example just using an Aspartate Aminotransferase [AST]-to-Platelet Ratio Index (APRI) score or ALT alone). The WHO guidance adopts a “treat-with” approach, where individuals with specified risk factors are offered treatment. Alternative strategies for determining treatment eligibility have been suggested, including a “treat-all-except” approach proposed by the HEPSANET consortium^7^, where all individuals are treated except those meeting criteria that suggest low risk of disease progression, and a “treat-all” approach, allowing same-day initiation of antiviral therapy in those who test positive for HBsAg without additional costly clinical staging^8^. Although effective at reducing HBV-associated disease burden, the clinical and public health implications and cost-effectiveness of a “treat-all” approach differs between settings^9–11^.

Liver fibrosis/cirrhosis staging is a recommended step in determining treatment eligibility. Since many individuals develop cirrhosis without clinical signs or symptoms, a clinical examination cannot be used to rule out compensated cirrhosis. Liver biopsy, the traditional gold standard, is invasive, costly, and impractical for routine use in many settings. Therefore, non-invasive methods, such as serum scoring systems and transient elastography (TE), are increasingly employed. TE, although effective (sensitivity ∼80% for detecting F2 fibrosis^12^), remains inaccessible to most patients due to the high costs of hardware purchase and maintenance, and limited availability^13^. The WHO guidelines recommend using the APRI (originally developed for use in chronic Hepatitis C infection^14^) as the preferred NIT. The Gamma-Glutamyl Transpeptidase-to-Platelet Ratio (GPR) has emerged as another NIT initially designed for HBV mono-infected individuals in a West African setting^15^.

A further consideration when designing HBV treatment strategies is understanding the prevalence of drug resistance within the population. Treatment guidelines recommend use of nucleos(t)ide analogues (NAs) that have a high genetic barrier to drug resistance (e.g. entecavir or tenofovir), and resistance is not widely regarded as a barrier to successful treatment either in individuals or at a public health level, as rates of acquired drug resistance (emergence of HBVDR due to viral replication and selection of RAMs in individuals exposed to NA therapy) are low for both agents in NA-naive individuals^16^. However, a systematic review found up to ∼20% entecavir resistance in NA-experienced individuals at 5 years^16^. Data is lacking from the WHO African region but suggests clinically relevant drug resistance may be present in some settings^17,18^. There is limited evidence to document the prevalence of transmitted drug resistance (HBV drug resistance in NA-naive people due to primary acquisition of infection with a virus with one or more RAMs), with current estimates varying widely by setting and drug agent, for example ranging from <1% to 37% in the WHO African region^17^.

This study aimed to assess treatment eligibility in a rural Ugandan population, using the new WHO guidelines. We investigated the sensitivity of NITs to predict liver fibrosis/cirrhosis, comparing sensitivity of APRI cut-offs in the WHO guidelines to HEPSANET thresholds and GPR thresholds for treatment. Finally we performed viral sequencing to determine the prevalence of drug resistance. By evaluating and comparing these approaches, this study aimed to contribute to discussions around efficient management of liver disease and improving health outcomes in resource-limited settings in sub-saharan Africa.

## 2. Methods

### 2.1 Uganda General Population Cohort and ULiDS sub-study

The MRC Uganda General Population Cohort (GPC) is a prospective population cohort that was established in 1989 to study HIV in rural south-western Uganda^19^. Since 2010, the scope of research questions investigated within the cohort has broadened to incorporate the epidemiology and genetics of both communicable and non-communicable diseases. The cohort now consists of ∼22,000 individuals living in 25 villages in the Kyamulibwa sub-county of the Kalungu district. All individuals were born prior to the introduction of childhood HBV vaccination (HepB3) under the Uganda National Expanded Programme on Immunization (UNEPI) in 2002.

In 2011 (GPC round 22), a baseline serosurvey for HIV, HBV and HCV infection was performed alongside liver function tests, lipid profile and assessment of cardiometabolic risk factors for 8099 individuals over the age of 13 years old (results previously reported^20^). This identified 220 people with positive Hepatitis B surface antigen (HBsAg) results, 81 of whom were re-surveyed in 2023, as part of the Uganda Liver Disease Study (ULiDS)^21^, with repeat liver function tests, full blood count, HBV serology, HBV VL, TE and HIV testing. Antiretroviral therapy with an HBV-active agent was available to those with HIV/HBV coinfection after HIV diagnosis but preceding this study, treatment for HBV monoinfection was not widely established.

### 2.2 Ethics

Ethics approval for the Uganda GPC was provided by the Science and Ethics Committee of the Uganda Virus Research Institute (GC/127/12/11/06 and GC/127/19/04/711), the Uganda National Council for Science and Technology (HS870), and the East of England-Cambridge South (formerly Cambridgeshire 4) NHS Research Ethics Committee UK (11/H0305/5); additional approval for the Uganda Liver Disease Study was provided by Oxford Tropical Research Ethics Committee (ref 50-18).

### 2.3 Laboratory assays

In 2011, liver function tests, HBV, HCV and HIV serology were performed as previously described^20^. In 2023, liver function tests were performed on the Roche cobas 6000 platform and full blood count on the BC 6200. A HBsAg rapid test was performed (Zhejiang Orient Gene Biotech Pal HBsAg rapid test) alongside HBV serology from a blood draw (Roche cobas 6000 platform, Elecsys HBsAg II, anti-HBc II (total IgM and IgG) and anti-HBs II assays). HIV testing was performed with the Abbott Determine HIV-1/2 rapid test. HBV VL levels were measured at the end of the study in stored samples collected in 2011 (59/220 available) and all individuals in 2023 using the Roche cobas Ampliprep/Taqman assay. Schistosoma mansoni infection is rare (prevalence of 1%) in this region and therefore was not tested for ^22^.

### 2.4 Non-invasive tests (NIT) for staging of liver disease

Non-invasive testing for liver disease was performed in 2023 with an APRI score (APRI = (AST level/40)/platelet count (10^9^/L) x100), GPR score (GPR = gamma-glutamyltransferase [GGT]/platelet count (10^9^/L)) and TE. APRI thresholds of 0.5^23^ and 0.36^24^ and a GPR cut-off of 0.32^15^ were used. Fasting TE was performed (Fibroscan^TM^, (Echosens Paris) with QC criteria of >= 10 measurements and IQR/Med < 30%) with cutoffs for significant liver fibrosis (>7kPa) and cirrhosis (>12.5kPa) were based on WHO 2024 CHB guidance. A description of Fibroscan^TM^ training events have been summarised in a report^25^.

### 2.5 Treatment criteria

We used the four treatment eligibility criteria set by the WHO in 2024 to determine treatment eligibility for all individuals with CHB^6^, the data used are summarised in **Table 1**. We then focused on individuals with HBV monoinfection, as those with HBV/HIV coinfection were already on HBV-active antivirals, to compare the sensitivity of different NITs for detecting liver fibrosis and cirrhosis.

**Table 1:** World Health Organisation HBV treatment guidance 2024. A summary of the treatment guidance and the parameters available in this cohort used to determine treatment eligibility.

| Criteria |  | Data available |
| --- | --- | --- |
| 1 | Treat all with significant fibrosis on non-invasive testing (APRI score >0.5 or transient elastography >7KPa), or clinical evidence of decompensated cirrhosis | - Clinical evidence of decompensated cirrhosis 2023 (ascites or jaundice)<br>- APRI 2023<br>- TE 2023 |
| 2 | HBV VL > 2000 IU/ml and an ALT level above the upper limit of normal (ULN) (30 U/L for men and boys and 19 U/L for women and girls) | - HBV VL and ALT in 2023 for all<br>- HBV VL and ALT in 2011 if available |
| 3 | Treat all with coinfections (such as HIV, hepatitis D or hepatitis C); family history of liver cancer or cirrhosis; comorbidities (such as diabetes or metabolic dysfunction–associated steatotic liver disease) and extrahepatic manifestations (e.g. glomerulonephritis or vasculitis) | - HIV 2011 and 2023<br>- HCV 2011<br>- Family history of death by liver disease<br>- Past medical history of diabetes or HbA1c >6.5 |
| 4 | Two ALT values above the ULN | - ALT in 2011 and 2023 |

ALT - Alanine Aminotransferase; APRI - Aspartate Aminotransferase-to-Platelet Ratio Index; TE - transient elastography, HbA1c - Hemoglobin A1c, HCV - hepatitis C virus, HIV - human immunodeficiency virus, ULN - upper limit of normal; VL - viral load

### 2.6 HBV whole genome sequencing and sequence analysis

HBV DNA was extracted from stored serum or plasma from 2011 and 2023 using the QIAamp MinElute Virus Spin Kit (Qiagen) with carrier RNA, as per the manufacturer’s protocol. For samples with VL>10^3^ IU/ml 200ul sample was used, and <10^3^ IU/ml, with 400ul, where sufficient volume was available. Whole genome sequencing was performed using the HEP-TILE tiled amplicon protocol and Oxford Nanopore Technology MinION^26,27^. Data were analysed using the hbv-fieldbioinformatics workflow^28^. HBV consensus sequences were used to determine genotypes, including recombinants, using the NCBI genotyping tool, using a pragmatic cut-off of >50% genome coverage^29^ and resistance associated mutations were determined using geno2pheno^30^.

### 2.7 Statistical analysis

Data analysis was performed using R v4.4.1^31^ using Fisher’s Exact Test to investigate the relationship between baseline HBeAg status and clinical parameters, ANOVA for parametric data, and Kruskal-Wallis test for non-parametric variables.

### 2.8 Data availability

Individual clinical data are not available in the public domain based on the governance restrictions for the General Population Cohort. The dataset used for this analysis will be archived by the London School of Hygiene and Tropical Medicine on publication in a peer reviewed journal. Sequences for samples where >50% genome coverage was generated were deposited to the European Nucleotide Archive (ENA) under the study numbers PRJEB79403 and PRJEB85529.

## 3. Results

### 3.1 Baseline characteristics and risk factors for liver disease

In 2011, 220/8099 (2.7%) individuals tested positive for HBsAg. On retrospective review of baseline and repeat HBV serology (HBsAg, anti-HBc, anti-HBs) and HBV VL, we identified ten individuals with false positive HBsAg results from 2011 and excluded these from further analysis, leaving 210 individuals with confirmed HBsAg (**Figure 1**). Baseline characteristics for the whole cohort and individuals with HBV infection are presented in **Table 2**. Among these 210, the median age was 29.5 years (IQR 19 to 42), 93/210 (44.3%) were female, and 24/210 (11.4%) were living with HIV/HBV co-infection. Self-reported alcohol intake was recorded for the whole cohort in 2011 where 2919 (36.0%) reported ever drinking alcohol. In 2023, 28/63 (44.4%) individuals with CHB reported drinking with daily alcohol intake reported by 5/63 (7.9%) and weekly by 14/63 (22.2%).

**Figure 1:**
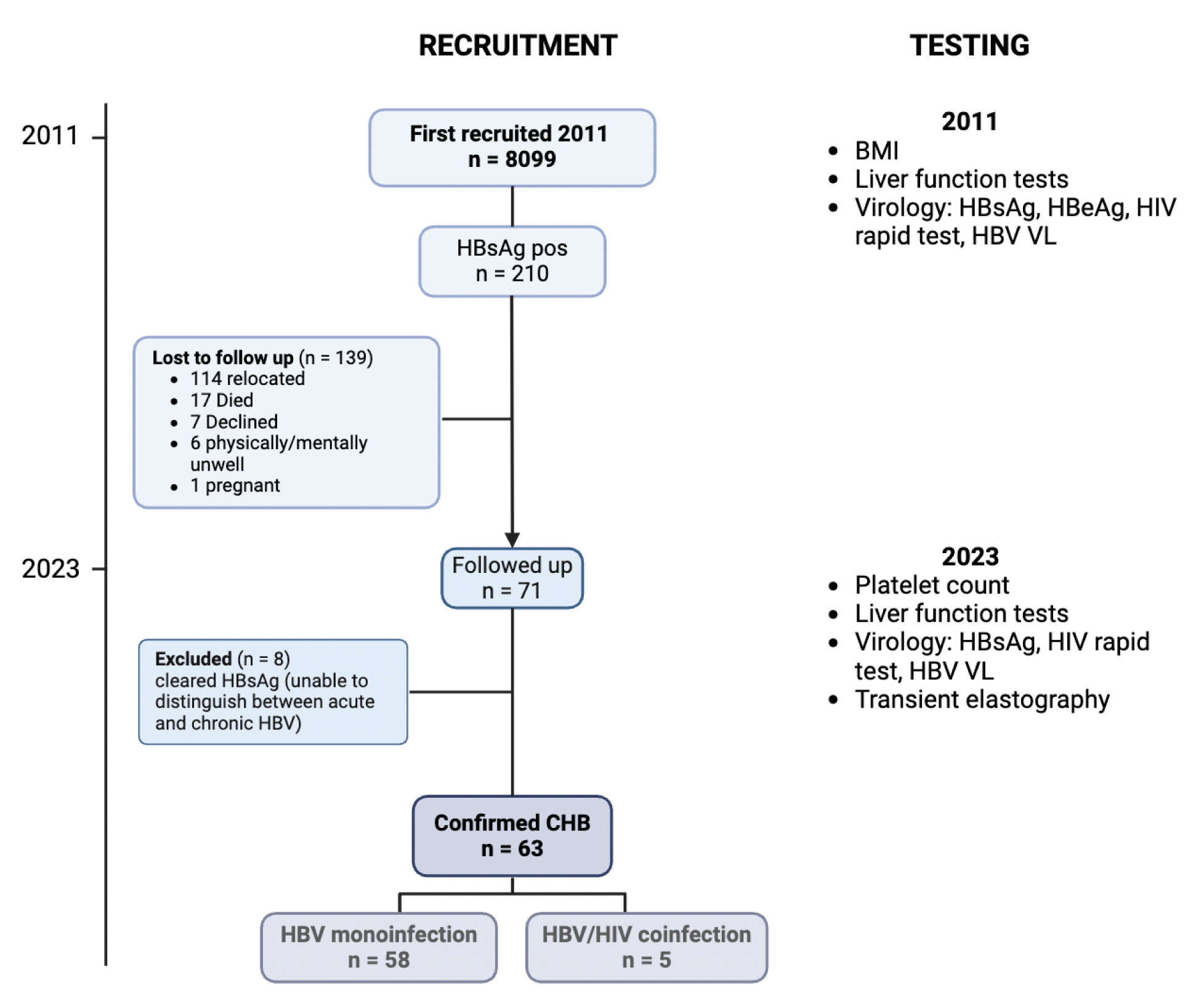
Flow chart showing the timeline for recruitment and testing of the Uganda liver disease (ULiDS) HBV cohort. Created in BioRender. Lumley, S. (2025) https://BioRender.com/d22b598 BMI - body mass index, CHB - chronic HBV, HBsAg - HBV surface antigen; HBeAg - HBV e antigen, HIV - human immunodeficiency virus, VL - viral load

**Table 2.**
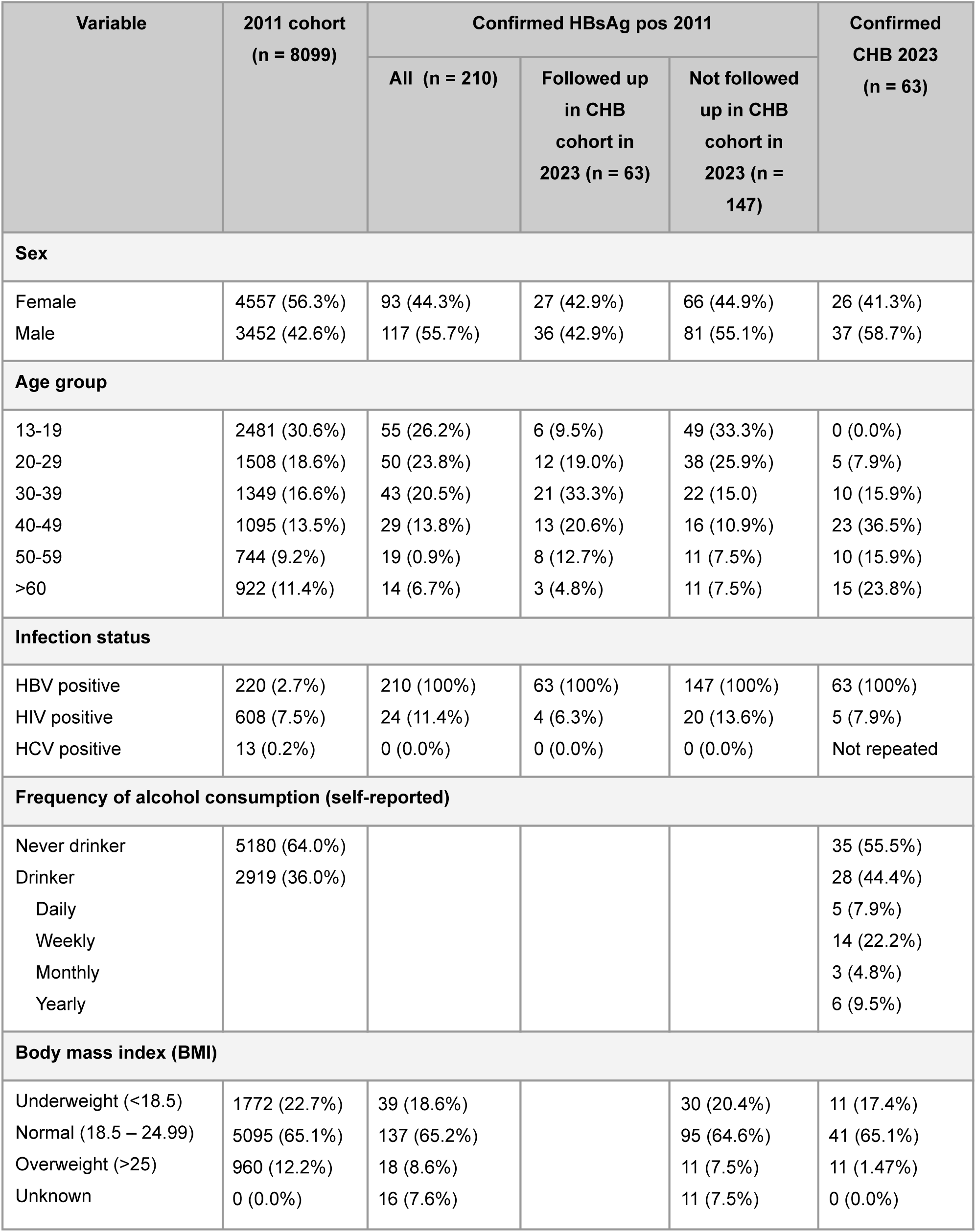
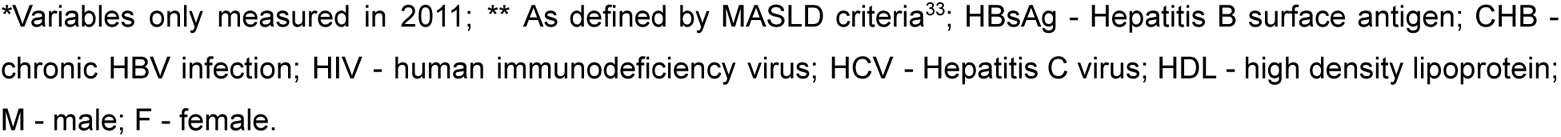
Uganda Liver Disease Study (ULiDS) cohort characteristics at baseline (2011) and in those followed up (2023)

| Variable | 2011 cohort<br>(n = 8099) | Confirmed HBsAg pos 2011 |  |  | Confirmed<br>CHB 2023<br>(n = 63) |
| --- | --- | --- | --- | --- | --- |
|  |  | All (n = 210) | Followed up<br>in CHB<br>cohort in<br>2023 (n = 63) | Not followed<br>up in CHB<br>cohort in<br>2023 (n = 147) |  |
| Sex |  |  |  |  |  |
| Female | 4557 (56.3%) | 93 (44.3%) | 27 (42.9%) | 66 (44.9%) | 26 (41.3%) |
| Male | 3452 (42.6%) | 117 (55.7%) | 36 (42.9%) | 81 (55.1%) | 37 (58.7%) |
| Age group |  |  |  |  |  |
| 13-19 | 2481 (30.6%) | 55 (26.2%) | 6 (9.5%) | 49 (33.3%) | 0 (0.0%) |
| 20-29 | 1508 (18.6%) | 50 (23.8%) | 12 (19.0%) | 38 (25.9%) | 5 (7.9%) |
| 30-39 | 1349 (16.6%) | 43 (20.5%) | 21 (33.3%) | 22 (15.0) | 10 (15.9%) |
| 40-49 | 1095 (13.5%) | 29 (13.8%) | 13 (20.6%) | 16 (10.9%) | 23 (36.5%) |
| 50-59 | 744 (9.2%) | 19 (0.9%) | 8 (12.7%) | 11 (7.5%) | 10 (15.9%) |
| >60 | 922 (11.4%) | 14 (6.7%) | 3 (4.8%) | 11 (7.5%) | 15 (23.8%) |
| Infection status |  |  |  |  |  |
| HBV positive | 220 (2.7%) | 210 (100%) | 63 (100%) | 147 (100%) | 63 (100%) |
| HIV positive | 608 (7.5%) | 24 (11.4%) | 4 (6.3%) | 20 (13.6%) | 5 (7.9%) |
| HCV positive | 13 (0.2%) | 0 (0.0%) | 0 (0.0%) | 0 (0.0%) | Not repeated |
| Frequency of alcohol consumption (self-reported) |  |  |  |  |  |
| Never drinker | 5180 (64.0%) |  |  |  | 35 (55.5%) |
| Drinker | 2919 (36.0%) |  |  |  | 28 (44.4%) |
| Daily |  |  |  |  | 5 (7.9%) |
| Weekly |  |  |  |  | 14 (22.2%) |
| Monthly |  |  |  |  | 3 (4.8%) |
| Yearly |  |  |  |  | 6 (9.5%) |
| Body mass index (BMI) |  |  |  |  |  |
| Underweight (<18.5) | 1772 (22.7%) | 39 (18.6%) |  | 30 (20.4%) | 11 (17.4%) |
| Normal (18.5 – 24.99) | 5095 (65.1%) | 137 (65.2%) |  | 95 (64.6%) | 41 (65.1%) |
| Overweight (>25) | 960 (12.2%) | 18 (8.6%) |  | 11 (7.5%) | 11 (1.47%) |
| Unknown | 0 (0.0%) | 16 (7.6%) |  | 11 (7.5%) | 0 (0.0%) |

\*Variables only measured in 2011; \*\* As defined by MASLD criteria<sup>33</sup>; HBsAg - Hepatitis B surface antigen; CHB - chronic HBV infection; HIV - human immunodeficiency virus; HCV - Hepatitis C virus; HDL - high density lipoprotein; M - male; F - female.

### 3.2 Characteristics at follow up in 2023

In 2023, 71/210 individuals with positive HBsAg were followed up (139/210 were lost to follow up, reasons detailed in **Figure 1**). In addition we identified eight individuals who were HBsAg negative and anti-HBc positive in 2023, having been HBsAg positive in 2011. These individuals are most likely to have cleared CHB, but may have had acute infection at the time of testing in 2011, so were excluded from further analysis in 2023. Characteristics of the remaining 63 individuals with confirmed CHB at follow up are detailed in **Table 2** and **Supplementary table 1**.

### 3.3 Eligibility for HBV treatment

Using the WHO 2024 HBV treatment guidelines (**Table 1**), we determined treatment eligibility in our cohort of 63 individuals living with CHB. In this group, a total of 24/63 (38%) individuals were eligible for treatment (**Figure 2**) based on meeting one or more of the four established treatment criteria (**Table 1**). The majority of individuals (19/24, 79%) were eligible on the basis of having ‘significant’ liver fibrosis (either APRI >0.5 or elastography > 7kPa), no individuals had evidence of decompensated cirrhosis (jaundice or ascites) on examination. Only one individual met the threshold for treatment based on criterion 2 (HBV VL >2000 IU/ml with ALT above upper limit of normal (ULN), see **Table 1** for definition); they had HBV VL 7 log_10_ IU/ml in 2011 which had fallen to 353 IU/ml in 2023 without treatment and had no evidence of fibrosis based on APRI or TE. Five individuals were eligible for treatment due to HIV coinfection (‘treat 3’), one of whom also met ‘treat 1’ fibrosis criteria. Only 1 individual met the ‘treat 4’ criteria of two ALT values above ULN.

**Figure 2:**
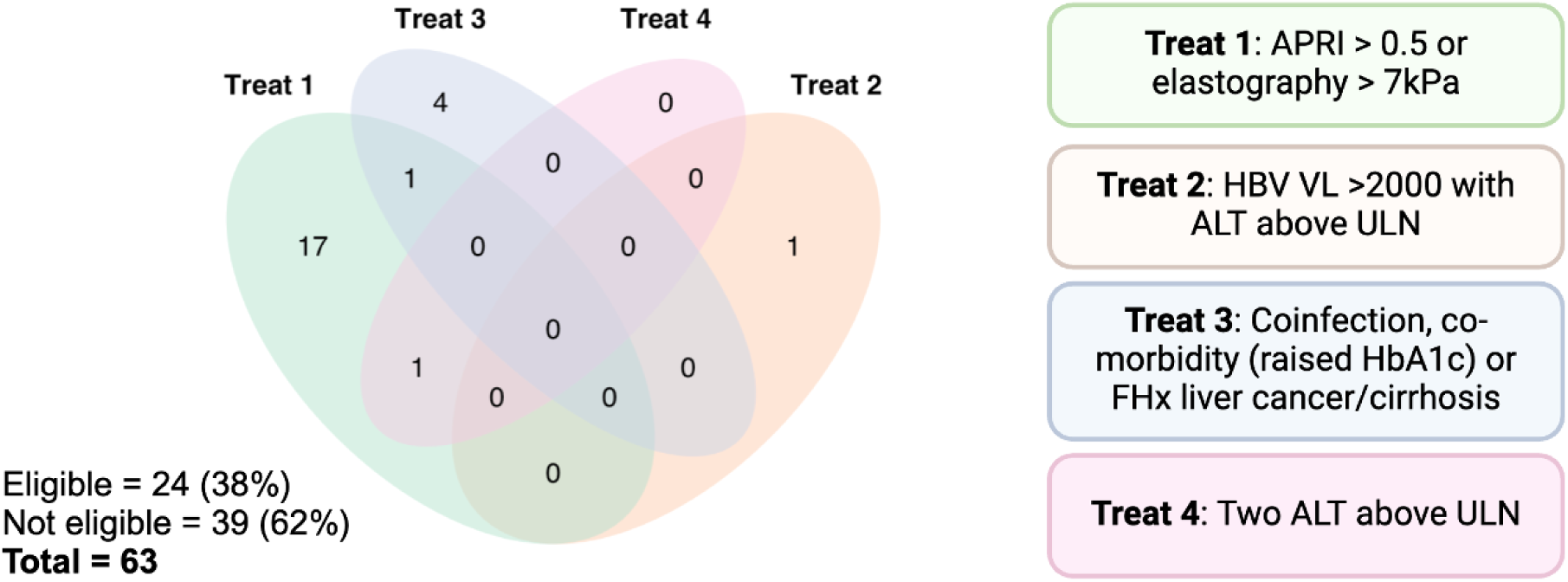
Eligibility for HBV treatment among 63 adults with chronic HBV in rural Uganda, based on WHO 2024 treatment guidelines. APRI - Aspartate Aminotransferase-to-Platelet Ratio Index, VL - viral load, ALT - alanine aminotransferase, ULN - upper limit of normal.

Given TE is not available in many clinical settings, we also determined treatment eligibility in a hypothetical scenario where TE was not available in this cohort. In this scenario only 14/63 (22%) individuals would meet treatment eligibility criteria.

### 3.4 Lack of concordance between non-invasive measures of liver disease in treatment-naive HBV mono-infected individuals

Using the WHO threshold of TE >7 kPa, 13/58 HBV mono-infected individuals were eligible for treatment. We investigated the sensitivity of different NIT for predicting fibrosis/cirrhosis in untreated individuals with CHB monoinfection, using TE as the gold standard.

#### 3.4.1 WHO 2024: “treat-with” APRI >0.5

Using a cut-off of APRI >0.5, 8/58 individuals were eligible for treatment; only 3 of whom had TE scores >7kPa. This APRI cut-off missed 10 individuals who would have been eligible for treatment using TE (**Figure 3A)**. In this cohort, APRI >0.5 had a low sensitivity for prediction of fibrosis and cirrhosis; an APRI threshold of >0.5 had a sensitivity of only 23.0% for predicting a TE score >7 kPa (F2 fibrosis) and 25.0% for predicting a TE score >12.5 kPa (F4 cirrhosis).

**Figure 3:**
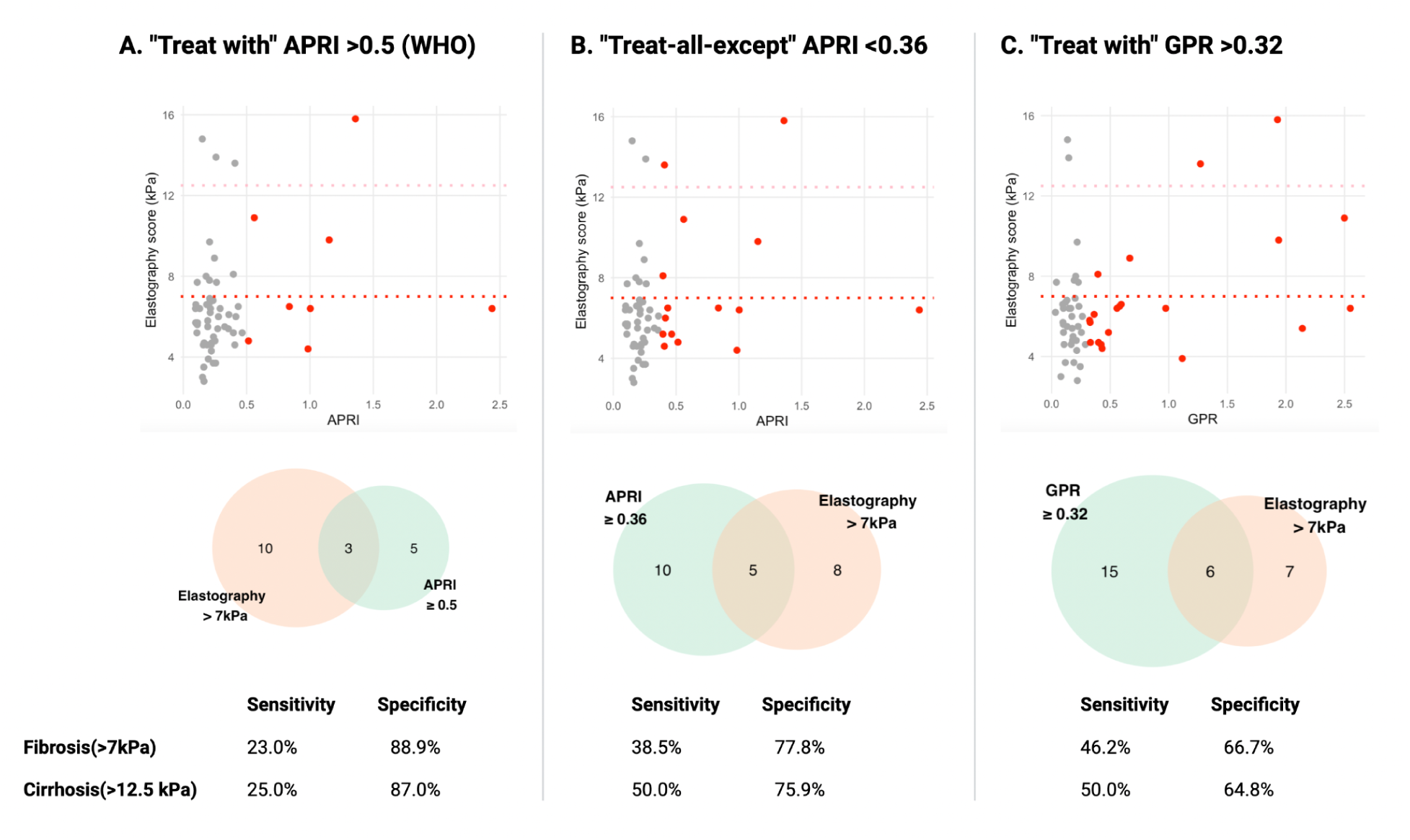
Predictive value of routinely available biomarkers for detecting fibrosis or cirrhosis in treatment-naive individuals with CHB mono-infection, using TE as the gold standard. Red dots indicate individuals with an APRI score >0.5, grey dots indicate those with an APRI score <0.5, red dotted line indicates TE score of 7kPa, pink dotted line indicates TE score of 12.5 kPa. WHO - World Health Organisation; APRI - Aspartate Aminotransferase-to-Platelet Ratio Index; GPR - Gamma-Glutamyl Transpeptidase-to-Platelet Ratio; TE - transient elastography

#### 3.4.2 HEPSANET: “treat-all-except” APRI <0.36

The ‘treat-all-except’ paradigm applies a more inclusive approach to treatment, where an optimised rule-out threshold of APRI <0.36 is used to exclude “low risk” individuals not requiring treatment, i.e. all individuals with APRI > 0.36 are offered treatment. Using this lower APRI cut-off, 15/58 individuals were eligible for treatment, with an increased sensitivity of 38.5% for detection of an elastography score >7 kPa (F2 fibrosis) and 50.0% for predicting an elastography score >12.5 kPa (F4 cirrhosis) (**Figure 3B**).

#### 3.4.3 PROLIFICA: “treat-with” GPR >0.32

Based on GPR >0.32, 21/58 individuals were eligible for treatment. Sensitivity for detecting fibrosis (>7kPa) was 46.2% and for cirrhosis (>12.5 kPa) was 50.0%. GPR >0.32 was the only fibrosis threshold met by the single individual meeting the “treat 2” criteria of HBV VL >2000 and ALT >ULN (GPR 0.324, APRI 0.19). (**Figure 3C**).

#### 3.4.4 “Treat-all”

If a ‘treat-all’ approach was used in this population, all 39/63 individuals who would have been ineligible based on all four of the assessment approaches recommended by WHO 2024 treatment guidelines would have received treatment, including all individuals with liver fibrosis/cirrhosis.

### 3.5 Which individuals are missed with biomarker score based NIT?

In this cohort, ten individuals with TE > 7kPa, who should have been eligible for treatment, were missed using APRI >0.5 alone. Only one of these individuals met one of the other treatment criteria, so 9/10 (90%) would have not received treatment in the absence of TE assessment. Two individuals with cirrhosis (TE >12.5 kPa) would not have met the threshold for treatment by any of the NITs or been captured by other WHO treatment criteria. One individual had a HBV VL of ∼90,000 IU/ml, the other ∼900 IU/ml, however both individuals had platelet count, ALT, AST and GGT within the reference range. Neither had co-infection with HIV or HCV and neither reported drinking alcohol. One individual had a BMI of 25.3 in 2011 (overweight) but had fallen to the normal range in 2023.

### 3.6 HBV VL as indication for treatment

Fifteen individuals had VL measured in both 2011 and 2023, 56 individuals in 2023 alone and 42 individuals in 2011 alone. The distribution of VL in HBV mono-infected untreated individuals is shown in **Figure 4**. All individuals with HIV coinfection on HBV-active ART had undetectable HBV VL in 2023, no other individuals were on antiviral treatment. Only one individual met the criteria for treatment based on HBV VL/ALT in 2011 or 2023 (HBV VL > 2000 IU/ml and an ALT >ULN).

**Figure 4:**
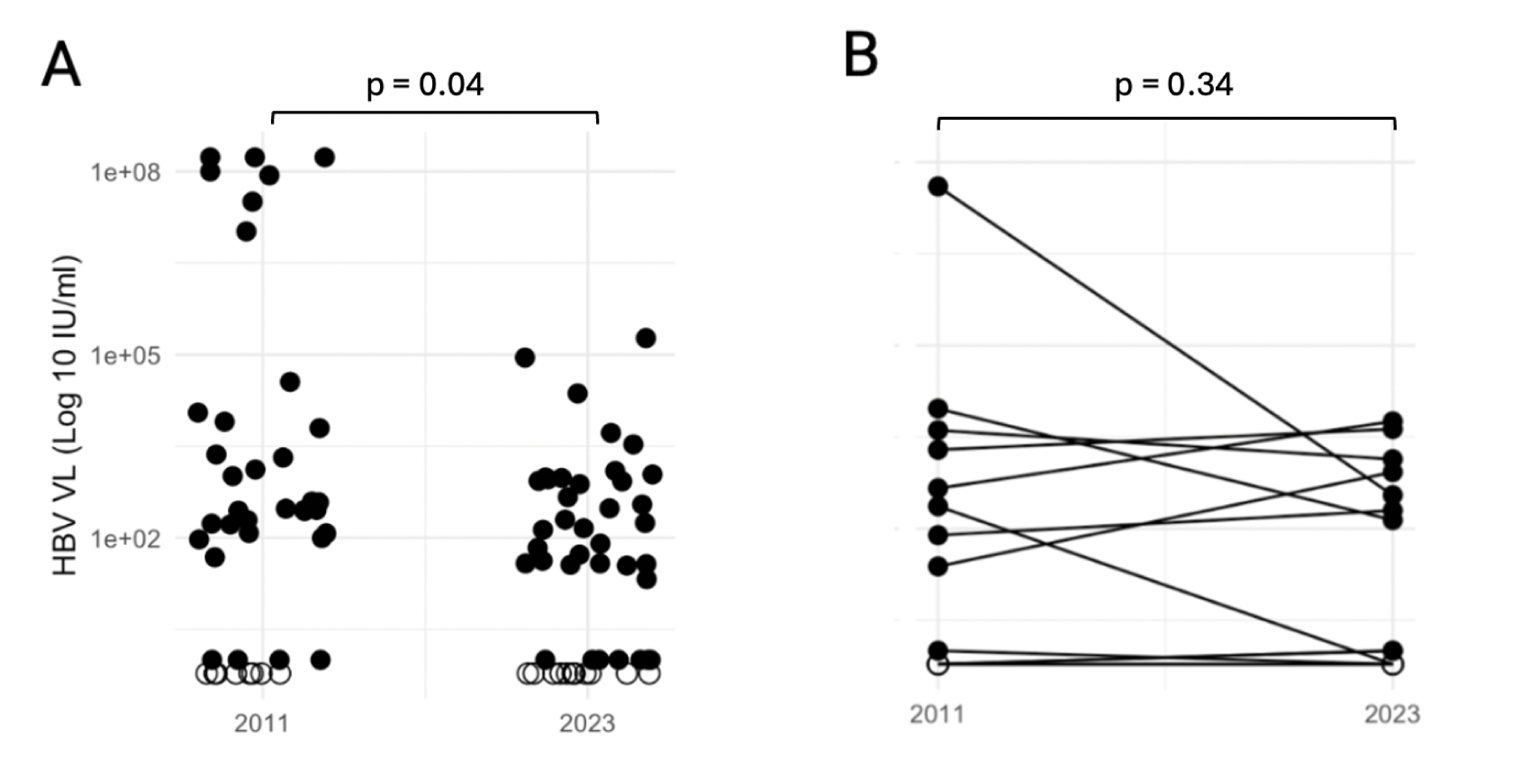
HBV VL in 2011 and 2023 in HBV mono-infected untreated individuals from the Uganda liver disease study (ULiDS). A) Individuals with HBV VL testing performed at a single time point (p = 0.04, Mann-Witney U test). B) Individuals with HBV VL testing performed at two time points (p = 0.34, paired t-test). Where an individual had samples at both time points, these are joined by a black line. Where HBV VL was undetected, the value is plotted as an empty circle (o), where HBV VL was detected but below the limit of quantification (<20 IU/ml), the value is plotted as 1.

### 3.6 Clinical correlations of HBV genotype

Eighty samples from seventy five individuals were available for sequencing (38 samples from 2011, 42 from 2023, and 5 individuals sequenced at both time points). Genotyping was performed for the 67 sequences with sufficient coverage; 57/67 were genotype A (subgenotype A1), 2/67 were A/D, 5/67 were D/E and 3/67 were A/D/E recombinants. There was no relationship between viral genotype and viral load, age, ALT, AST, APRI score, GPR score or TE score, however numbers of non A genotypes were very small.

### 3.7 HBV Drug Resistance (HBVDR)

We looked for evidence of HBVDR in HBV mono-infected individuals. In this cohort, 37/58 individuals with HBV mono-infection had quantifiable HBV VL among whom 29 had virus sequenced. No evidence of HBVDR was seen.

In this cohort, 10/24 individuals with HIV/HBV coinfection had retrospective HBV VL performed on biobank samples from 2011; 6/10 individuals had quantifiable HBV VL, 5/10 had virus sequenced. Of these, 2/5 had the RAM combination rt180M/204V (both genotype A with HBV VL 80-90 million IU/ml).

There was no evidence of HBVDR in the five individuals with HIV/HBV coinfection in the CHB cohort followed up in 2023; all individuals were virologically suppressed on HBV-active ART.

## 4. Discussion

We applied the 2024 WHO HBV treatment criteria to a cohort of individuals with CHB in rural Uganda, establishing the percentage of individuals eligible for treatment (38%). We highlight how treatment eligibility varies according to the eligibility criteria and scoring system used. Diagnostic availability impacts on ability to identify individuals who might benefit from treatment.

When implementing the new WHO HBV guidelines in settings without reliable access to TE or HBV VL, NITs of liver disease such as APRI are applied as second-line approaches for assessing treatment eligibility. In this population, 24/63 (38%) were eligible for treatment using all data available (**Table 1**), this fell to 14/63 (23%) in a hypothetical scenario in which TE was not available and APRI >0.5 was relied on to identify those with liver fibrosis and cirrhosis. Our findings raise concerns about the adequacy of the APRI threshold >0.5 as a predictor of liver fibrosis and cirrhosis in this population, where this APRI threshold fails to identify the majority of individuals with fibrosis and cirrhosis (sensitivity 23% and 25% respectively) and leads to missed opportunities for treatment. The sensitivity we see contrasts to that referenced in the WHO HBV guidelines, in which an APRI >0.5 was associated with a sensitivity of 71.7% (95% CI 67.1–75.8%) for identifying ≥F2 among people with CHB infection, although the guidelines acknowledge a lack of evidence from the WHO African region^5^. The possibility of false negative results is raised in the WHO guidance, and mitigated against by saying that people with CHB infection may meet other eligibility criteria for antiviral therapy (such as elevated HBV VL). However, in this cohort, only 10% of individuals with a false negative APRI score met another treatment criterion. We demonstrate improved, but still limited, sensitivity using a ‘treat-all-except’ threshold of APRI <0.36 or a ‘treat-with’ threshold of GPR >0.32, and the increase in sensitivity is coupled with a fall in specificity.

In this cohort, a combination of elevated ALT/HBV VL (‘treat 2’) and a persistently elevated ALT (‘treat 4’) identified one individual each for treatment (1.6%). This contrasts with the WHO estimates that ALT/HBV VL would capture an estimated 20-35% of all HBsAg positive people and that persistently positive ALT would capture 20%. Relative to the expectation that 50% of most CHB populations would be treatment eligible based on new WHO guidelines^32^, the relatively lower eligibility in this cohort could be driven by the older age at follow up, associated with HBeAg negative status and lower HBV VL. Furthermore, there is a low frequency of ALT and HBV VL testing compared to the recommended 12 monthly tests; this reduces the chance of individuals meeting the ‘treat 4’ criterion. Despite these caveats, these data raise the question about the incremental benefit of performing HBV VL in this population for the purposes of treatment stratification. HBV VL testing is not readily available in many settings, and where available, it is expensive either for state-funded health services or for the individual paying out-of-pocket.

Together, the lack of access and expense of TE (the gold standard NIT), the poor sensitivity of cheaper, accessible NITs such as APRI, and the low contribution and high cost of HBV VL raise some fundamental questions. Is it preferable to risk over-diagnosis or under-diagnosis of liver fibrosis/cirrhosis in this and similar settings? While over-diagnosis might increase unnecessary treatment and increase drug costs, under-diagnosis risks failing to offer timely interventions to prevent further morbidity and mortality. In resource limited settings, without appropriate risk stratification tools, should we be treating all HBsAg positive individuals? A treat-all approach would effectively reduce HBV-associated morbidity and mortality, reduce the cost of evaluation, overcome challenges with shifting eligibility over time, and negate the issue that different NITs identify different sets of individuals for treatment. Furthermore it would provide a more effective suppression of the HBV reservoir by reducing viraemia in individuals not otherwise eligible for treatment (a benefit not included in current cost estimates^9^). However, acceptability, the burden of life-long treatment and economic feasibility remain concerns^9^. In this cohort, all individuals eligible for treatment based on the WHO 2024 guideline will be offered treatment.

The absence of HBVDR in the HBV mono-infected population was reassuring. The presence of rt180M/204V in two individuals with HIV-HBV coinfection may be related to prior lamivudine exposure, however drug histories were not available. The combination rt180M/204V causes lamivudine resistance in HBV, and reduced susceptibility to entecavir and adefovir, and is also part of a combination of mutations recognised in association with tenofovir resistance. We did not observe a relationship between genotype and clinical outcomes, however the number of individuals with non-A genotype was small.

We acknowledge this study has limitations. The high loss to follow-up, particularly due to migration of younger individuals, presents challenges for the assessment of treatment eligibility and health outcomes. Although TE is accepted globally as an alternative to liver biopsy for the identification of liver fibrosis and cirrhosis in CHB, TE (as with all tests) has limitations in sensitivity and specificity (sensitivity using a cut-off of >6.0 to 8.0 kPa for F2 fibrosis 75.1% (95% CI 72.2–77.7%), specificity 79.3% (95% CI 76.2–82.2%) compared to liver biopsy which itself has issues with inter-reporter reliability and sampling error^5^). Individuals were asked to fast prior to testing in this cohort, however false positive TE scores may occur if individuals were not fasting, or due to liver inflammation for other reasons (eg alcohol consumption, other intercurrent infections). People with hepatitis flares or acute hepatitis will have falsely high liver stiffness measurements, however no individuals with high TE and low APRI had raised liver ALT or AST. In a clinical setting a repeat TE may be performed at a later date to clarify the clinical picture, however this was not possible within the timeframe of this study. We planned to evaluate the TREAT-B score (based on ALT and HBeAg status) however HBeAg testing was only performed on part of the cohort in 2011 and not available in 2023, highlighting issues with assay availability even in established research settings.

In conclusion, the WHO 2024 HBV treatment guidelines make important progress in facilitating access to treatment for people living with CHB. However, the current lack of accurate, low-cost diagnostics to identify individuals with liver fibrosis and cirrhosis raises concerns that these criteria may miss individuals who would benefit from intervention, particularly in settings where TE is not available. Our study emphasizes the need for longitudinal research in Africa to clarify which individuals benefit most from treatment, which biomarkers are the most appropriate predictors of outcome, the practicality of different approaches, and their affordability. WHO guidelines also highlight the urgent need for contextually appropriate diagnostic strategies tailored to the realities of healthcare delivery in African populations. Addressing these challenges is vital for improving outcomes in the management of CHB and achieving global health equity.

## ACKNOWLEDGEMENTS

We thank all the participants in the Uganda General Population Cohort and all the staff at the MRC/UVRI and LSHTM Uganda Research Unit Kyamulibwa office for supporting our work.

---

**Supplementary Table 1.** Uganda Liver Disease Study (ULiDS) chronic HBV cohort: viral load, ALT, APRI score and transient elastography score at follow up in 2023.

| Variable | Baseline HBeAg status |  |  | Overall<br>(n = 63) | P-value* |
| --- | --- | --- | --- | --- | --- |
|  | Baseline HBeAg<br>positive<br>(n = 9) | Baseline HBeAg<br>negative<br>(n = 37) | Baseline HBeAg<br>unknown<br>(n = 17) |  |  |
| HBV viral load (IU/ml) |  |  |  |  |  |
| < 2000 | 6 (66.7%) | 35 (95.0%) | 15 (88.0%) | 56 (89.0%) | 0.02 |
| 2000-20,000 | 1 (11.0%) | 2 (5.4%) | 1 (5.9%) | 4 (6.3%) |  |
| > 20,000 | 2 (22.0%) | 0 (0.0%) | 1 (5.9%) | 3 (4.8%) |  |
| Alanine amino transferase (ALT) |  |  |  |  |  |
| Normal | 8 (89%) | 35 (95%) | 16 (94%) | 59 (94%) | 0.49 |
| Raised** | 1 (11%) | 2 (5.4%) | 1 (5.9%) | 4 (6.3%) |  |
| APRI score |  |  |  |  |  |
| > 1 (F4 cirrhosis) | 1 (11%) | 2 (5.4%) | 1 (5.9%) | 4 (6.3%) | 0.08 |
| 0.5 - 1 (F2 fibrosis) | 2 (22%) | 1 (2.7%) | 1 (5.9%) | 4 (6.3%) |  |
| < 0.5 (Normal) | 6 (67%) | 34 (92%) | 15 (88%) | 55 (87.0%) |  |
| Transient elastography score (kPa) |  |  |  |  |  |
| > 12.5 (F4 cirrhosis) | 2 (22%) | 1 (2.7%) | 1 (5.9%) | 4 (6.3%) | 0.02 |
| 7 - 12.5 (F2 fibrosis) | 3 (33%) | 4 (11%) | 3 (18.0%) | 10 (16.0%) |  |
| < 7 (Normal) | 4 (44%) | 32 (86%) | 13 (76.0%) | 49 (78.0%) |  |

\* p-value comparing those with known HBeAg status (Fisher's Exact Test)
\*\* Above 30 U/L for men and boys and 19 U/L for women and girls

